# Stratification of COVID-19 severity using SeptiCyte RAPID, a novel host immune response test

**DOI:** 10.1101/2022.09.15.22279735

**Authors:** Victor Gravrand, François Mellot, Felix Ackermann, Marie-Christine Ballester, Benjamin Zuber, James T. Kirk, Krupa Navalkar, Thomas D. Yager, Fabien Petit, Tiffany Pascreau, Eric Farfour, Marc Vasse

**Affiliations:** Biology Department, Foch Hospital, Suresnes, France; Radiology Department, Foch Hospital, Suresnes, France; Internal Medicine Department, Foch Hospital, Suresnes, France; Emergency Department, Foch Hospital, Suresnes, France; Intensive Care Unit, Foch Hospital, Suresnes, France; Immunexpress, Inc., Seattle, Washington, United States); UMR-S1176, Le Kremlin-Bicêtre, France

**Keywords:** COVID-19, gene expression, severity, viral sepsis

## Abstract

SeptiCyte^®^ RAPID is a gene expression assay measuring the relative expression levels of host response genes PLA2G7 and PLAC8, indicative of a dysregulated immune response during sepsis. As severe forms of COVID-19 may be considered viral sepsis, we evaluated SeptiCyte RAPID in a series of 94 patients admitted to Foch Hospital (Suresnes, France) with proven SARS-CoV-2 infection. EDTA blood was collected prospectively in the emergency department (ED) in 67 cases, in the intensive care unit (ICU) in 23 cases and in conventional units in 4 cases. SeptiScore (0-15 scale) increased with COVID-19 severity. Patients in ICU had the highest SeptiScores, producing values comparable to 8 patients with culture-confirmed bacterial sepsis. Receiver operating characteristic (ROC) curve analysis had an area under the curve (AUC) of 0.81 for discriminating patients requiring ICU admission from patients who were immediately discharged or from patients requiring hospitalization in conventional units. SeptiScores increased with the extent of the lung injury. For 68 patients, a chest computed tomography (CT) scan was performed within 24 hours of COVID-19 diagnosis. SeptiScore > 7 suggested lung injury ≥ 50 % (AUC = 0.86). SeptiCyte RAPID was compared to other biomarkers for discriminating Critical + Severe COVID-19 in ICU, versus Moderate + Mild COVID-19 not in ICU. The mean AUC for SeptiCyte RAPID was superior to that of any individual biomarker or combination thereof. In contrast to C-reactive protein (CRP), correlation of SeptiScore with lung injury was not impacted by treatment with anti-inflammatory agents. SeptiCyte RAPID can be a useful tool to identify patients with severe forms of COVID-19 in ED, as well as during follow-up.

## Introduction

Sepsis is a complex immuno-pathological disorder characterized by a pro-inflammatory response that may be followed by an anti-inflammatory, immunosuppressive state, or possibly a cycling between these states over several weeks [1]. Sepsis-3 now defines sepsis as “a life-threatening organ dysfunction caused by a dysregulated host immune response to infection”. It is now well known that SARS-CoV-2 infection can progress to a severe disease characterized by a dysregulated immune response and a maladaptive cytokine release [2], leading to multiple complications including acute respiratory distress syndrome (ARDS), myocarditis, acute kidney and liver failure and coagulopathy [3].Therefore there are important similarities of this severe disease progression to sepsis. As such, Beltran-Garcia et al. [4] reviewed the common features of COVID-19 and sepsis and discussed the possible use of anti-inflammatory therapeutics in the treatment of SARS-CoV-2 disease. In a large meta-analysis of 24,983 patients with COVID-19, Abate et al. [5] reported that approximately one third of these patients were admitted to the Intensive Care Unit (ICU) and 39% of those patients died.

Since the start of the COVID-19 pandemic in 2020, the disease has evolved in several waves, corresponding to different variants, and is characterized by rapid and massive contamination of patients, who arrive in large numbers in the emergency department (ED). Severity of the disease is evaluated by different biological parameters such lymphopenia, blood oxygen saturation, C-reactive protein (CRP) levels and D-dimer levels [6]. In addition, chest computed tomography (CT) scan has been proposed to identify patients at risk of poor evolution [7]. Because of huge influxes of patients at the same time in the ED, laboratory and radiology departments can be overwhelmed, due to this massive activity and possibly the absence of medical workers, themselves affected by the disease, leading to a delay in the triage of the patients. Therefore, a test that would be able to determine easily and quickly the clinical severity would be welcome.

SeptiCyte^®^ RAPID (Immunexpress, Seattle, WA) is a gene expression assay using reverse transcription quantitative polymerase chain reaction (RT-qPCR) to measure the relative expression levels of host response genes (PLA2G7 and PLAC8) that are indicative of a dysregulated immune response during sepsis. SeptiCyte RAPID is used in conjunction with clinical assessments, vital signs and laboratory findings as an aid to differentiate infection-positive (sepsis) from infection-negative systemic inflammation in patients suspected of sepsis [8]. SeptiCyte RAPID generates a score (SeptiScore^®^) that falls within discrete Interpretation bands based on the increasing likelihood of infection-positive systemic inflammation. Analysis of public next generation sequencing (NGS) datasets (e.g. [9]) suggested that SeptiCyte RAPID is able to discriminate between milder and more severe COVID-19 cases.

Here, we report on a prospective study that sought to evaluate the performance of the SeptiCyte RAPID assay in patients with laboratory-confirmed COVID-19 with different degrees of severity, and analyze whether this assay could be used as a triaging tool for patients requiring hospitalization and potentially ICU care. A preliminary version of this work has been presented in abstract form [10].

## Methods

### Ethic committee approval

This study was conducted under a blanket protocol for COVID-related research at the Foch Hospital (“Information réutilisation des données personnelles et des échantillons biologiques à des fins de recherche V2 - 9 avril 2020”), which had been established and approved by the local ethics committee of the Foch Hospital (IRB00012437). The study was performed in agreement with French ethical laws, such that the patients and their families were informed that their biological samples and data used for routine care could be used in an anonymous manner unless they expressed their opposition. No patients enrolled in this study, or their families, expressed opposition to the use of their biological samples and data, which according to the French ethical laws and Foch Hospital ethics committee constitutes informed consent.

### Patient recruitment

The study was performed over a six month prospective recruitment period between mid-January and mid-July, 2021, when the SARS-CoV-2 B.1.1.7 variant was present in France. Ninety-four patients (44 females, 50 males, median age 65 years) were included in the study. All enrolled patients were confirmed by RT-qPCR to be nasopharyngeal swab positive for SARS-CoV-2 infection. Figure 1 presents a diagram of the flow of the patients in this study through the ED, conventional units and ICU, indicating the hospital locations where blood sampling for SeptiCyte measurement occurred. A ‘conventional unit’ is defined as a general hospital ward, with extra precautions and isolations taken to minimize SARS-CoV-2 transmission.

**Figure 1.**
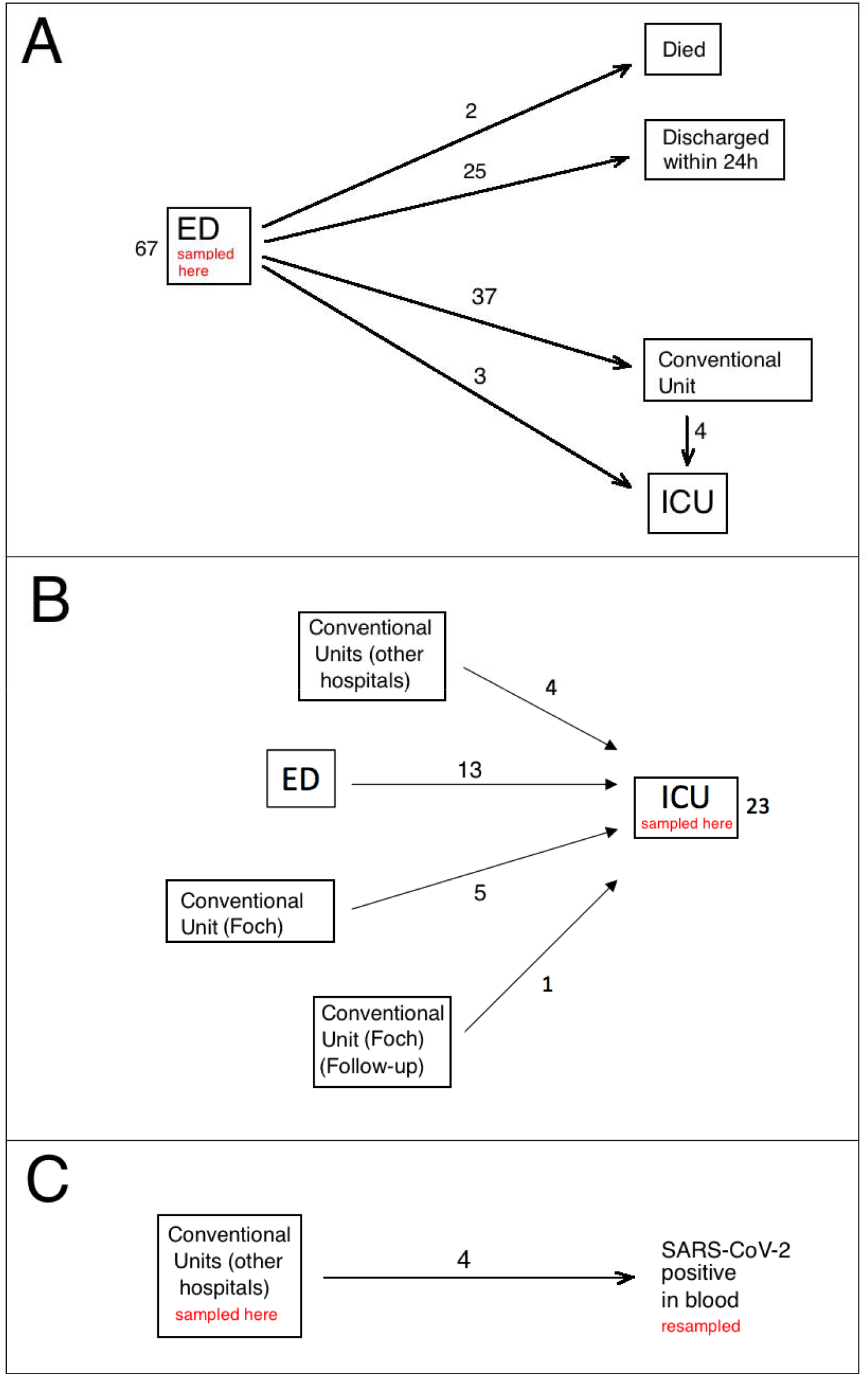
Diagram of the flow of the patients in this study. (A) Sixty-seven patients sampled in ED. (B) Twenty-three patients sampled in ICU. (C) Four patients sampled by nasopharyngeal swab in the conventional unit, with subsequent SARS-CoV-2 (+) result from blood.

### Collection of blood samples

K3-EDTA whole blood samples, collected for complete blood count (CBC) and subsequently tested with SeptiCyte RAPID, were collected on admission in the ED in 67 cases (Fig. 1A), in ICU for 23 patients (Fig. 1B) and in conventional units for 4 patients (Fig. 1C). Of the 67 patients tested in the ED, 25 were discharged within 24 hours, because they had a moderate clinical form, did not require oxygen therapy, and had no biological signs of marked inflammation. None of these 25 patients required another hospitalization for COVID-19 and all were alive at least one month after their admission to the ED. Thirty-three patients were transferred from ED to conventional units for a median time of 8 days (range 2 to 22 days), four were first transferred from ED to conventional units, but then required admission to ICU some days later (median time 6 days, range 1 -12 days), and three were immediately transferred from ED to ICU. Two older patients (>80 years of age) died in the ED.

Twenty-three patients were tested in ICU (fig 1B). Of these, 13 were admitted first to the ED (within 24 hours) but required high levels (>15L/min) of oxygenotherapy and therefore were transferred to ICU as soon as possible, with nasopharyngeal swab RT-qPCR and blood sample collection occurring after ICU admission. Another group of 9 patients in ICU came from conventional units, either from Foch hospital (5 cases) or from other hospitals (4 cases). Median length of stay in conventional units before ICU admission was 3 days (range 2 to 5 days). For the last case (labeled “Follow-up” in Fig. 1B), the SeptiScore was determined during ICU stay, because the patient was tested during a follow-up for COVID-19 by RT-qPCR, and was still positive 35 days after the initial diagnosis of COVID-19. Of the 23 patients tested in ICU (Fig. 1B), 22/23 were treated by corticotherapy and 11/23 were treated by antibiotherapy at the time of the SeptiScore determination. A blood culture was taken on the day SeptiCyte RAPID testing was performed in 20 cases, and was positive in 2 cases (*Klebsiella pneumoniae* and *Pseudomonas aeruginosa*). Similarly, cytobacteriological examination of the urine was taken from 12 patients, and was positive in 2 cases (*Klebsiella pneumoniae* and *Enterobacter cloacae*). On the day of SeptiCyte RAPID testing, the SOFA score was available for 20 patients and the median value was 2 (range: 0 – 12). For 10 ICU patients, follow-up samples were analyzed during ICU hospitalization, spaced approximately one week apart.

Lastly (Fig. 1C), for 4 patients hospitalized in conventional units, a sample was analyzed during their hospitalization (median time: 7 days, range: 3 – 15 days). These patients were initially hospitalized because they had a positive RT-qPCR on nasopharyngeal swab, but on the day when the SeptiScore was analyzed, SARS-CoV-2 viral sequences were also found in their blood. Testing for SARS-CoV-2 in blood samples was not performed for all the patients, but only for patients in whom the use of convalescent plasma was envisaged.

For comparison, we also quantified the SeptiScore in 8 patients from ICU with culture proven bacterial sepsis as per the Sepsis-3 criteria [11], who were SARS-CoV-2 negative.

### Processing of samples

Diagnosis of SARS-CoV-2 infection was made using either the Alinity M SARS-CoV-2 RT-PCR assay (Abbott Molecular, Des Plaines, IL, USA) or the Xpert^®^ Xpress SARS-CoV-2 (Cepheid Europe SAS, Maurens-Scopont, France)

Complete blood counts (CBC) were performed on a DxH 800 instrument (Beckman-Coulter, Villepinte, France) and monocyte CD16 positive quantification was performed using the CytoDiff™ reagent (Beckman-Coulter) as previously described [12]. Blood gas analysis, including lactate measurement, was performed on a GEM 4000 instrument (Werfen, Le Pré-Saint-Gervais, France). CRP and D-dimer were measured on Alinity IC (Abbott Diagnostics, Rungis, France) and ACL 700 (Werfen) systems, respectively. All measurements were made at approximately the same time the blood samples were collected for CBC and SeptiCyte RAPID testing.

SeptiCyte RAPID testing of eligible patients was performed on qualified Biocartis Idylla instruments (Biocartis, Mechelen, Belgium) installed with SeptiCyte RAPID assay software. Residual EDTA blood samples from CBC were diluted within several hours of collection with phosphate buffered saline (PBS; 1 vol blood to 2.75 vol PBS), and 900 μL of each diluted sample was put into a SeptiCyte RAPID cartridge and automatically processed.

The SeptiCyte RAPID test is completely self-contained within single use cartridges that are designed to run on the Biocartis Idylla system. The entire assay process from blood sample extraction to final test result is achieved on-cartridge, with fully integrated sample preparation followed by reverse transcription and quantitative real-time polymerase chain reaction (RT-qPCR) for the detection and quantification of changes in expression of the immune host response genes PLAC8 and PLA2G7.

The test result (SeptiScore) is calculated based on the difference of cycle of quantification (Cq) values in RT-qPCR for the target genes. SeptiScores range from 0-15 and higher scores are associated with increasing sepsis probabilities. A sample processing control (SPC) is included in each cartridge to confirm that adequate nucleic acid extraction and amplification has occurred.

### Radiologic evaluation

COVID-19 disease severity was assessed as the percentage of specific lung abnormalities or damage as observed by a radiologist on a chest CT scan: (1) peripheral, bilateral ground glass opacity (GGO) with or without consolidation or visible intra-lobular lines (crazy paving), or (2) multifocal GGO of rounded shape with or without consolidation or visible intra-lobular lines. Patients were then classified as “absence”, “mild”, “moderate”, “extensive”, “severe”, or “critical” based on 0%, present but <10%, 10-25%, 25-50%, 50-75%, or >75% lung damage respectively. Three patients were indicated as having “non-specific” CT scan results, and were not considered further in the classification totals.

### Statistical analyses

Statistical analyses were performed using a combination of publicly available and custom scripts written in the R programming language [13]. The pROC package in R [14] was used for AUC calculations, and some calculations were verified with JLABROC4 [www.rad.jhmi.edu/jeng/javarad/roc/JROCFITi.html]. The stats package in R was used for calculating p-values with Welch’s two-sided t-test. One-way ANOVA was conducted using the gtsummary package in R.

Combinatorial regression analysis, in which values of multiple clinical parameters (SeptiScore, CRP, D-dimer, lactate, monocytes, CD16-positive monocytes) were combined, was performed with a custom R script. Prior to the combinatorial analysis, the R package Amelia [15] was used to impute missing values in the clinical dataset. (The numbers of missing values were 1 for monocytes; 7 for CD16-monocytes, 9 for D-dimer; 11 for lactate; and 1 for CRP.)

In box-and-whisker plots (Figures 2, 3) the following features were defined: lower edge, midline, and upper edge of box: 25^th^, 50^th^, 75^th^ percentiles respectively; lower whisker boundary: smallest observed value above the 25th percentile - 1.5 * IQR; upper whisker boundary: largest observed below the 75th percentile + 1.5 * IQR.

**Figure 2.**
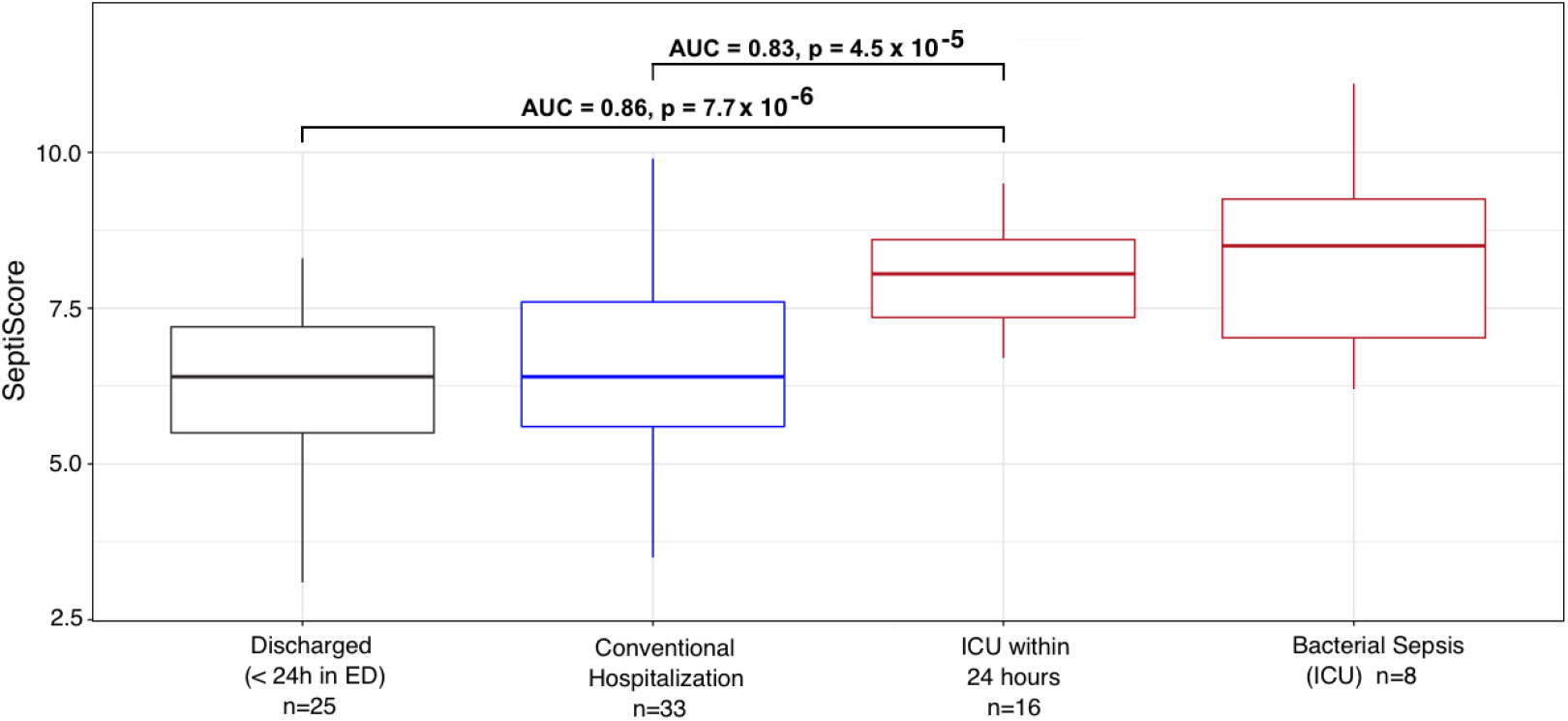
Variation of the SeptiScore in COVID-19 patients determined within 24 hours after hospital admission according to the location of the patients. Comparison with SeptiScores of patients with culture-confirmed bacterial sepsis. COVID patients: Group 1 – Discharged within 24 hours (n = 25, Fig 1a); Group 2 – Hospitalization in conventional unit, without subsequent admission to ICU (n = 33, Fig 1a); Group 3 – transferred to ICU within 24 hours (n= 16); Group 4 – Bacterial sepsis in ICU (n = 8).

**Figure 3.**
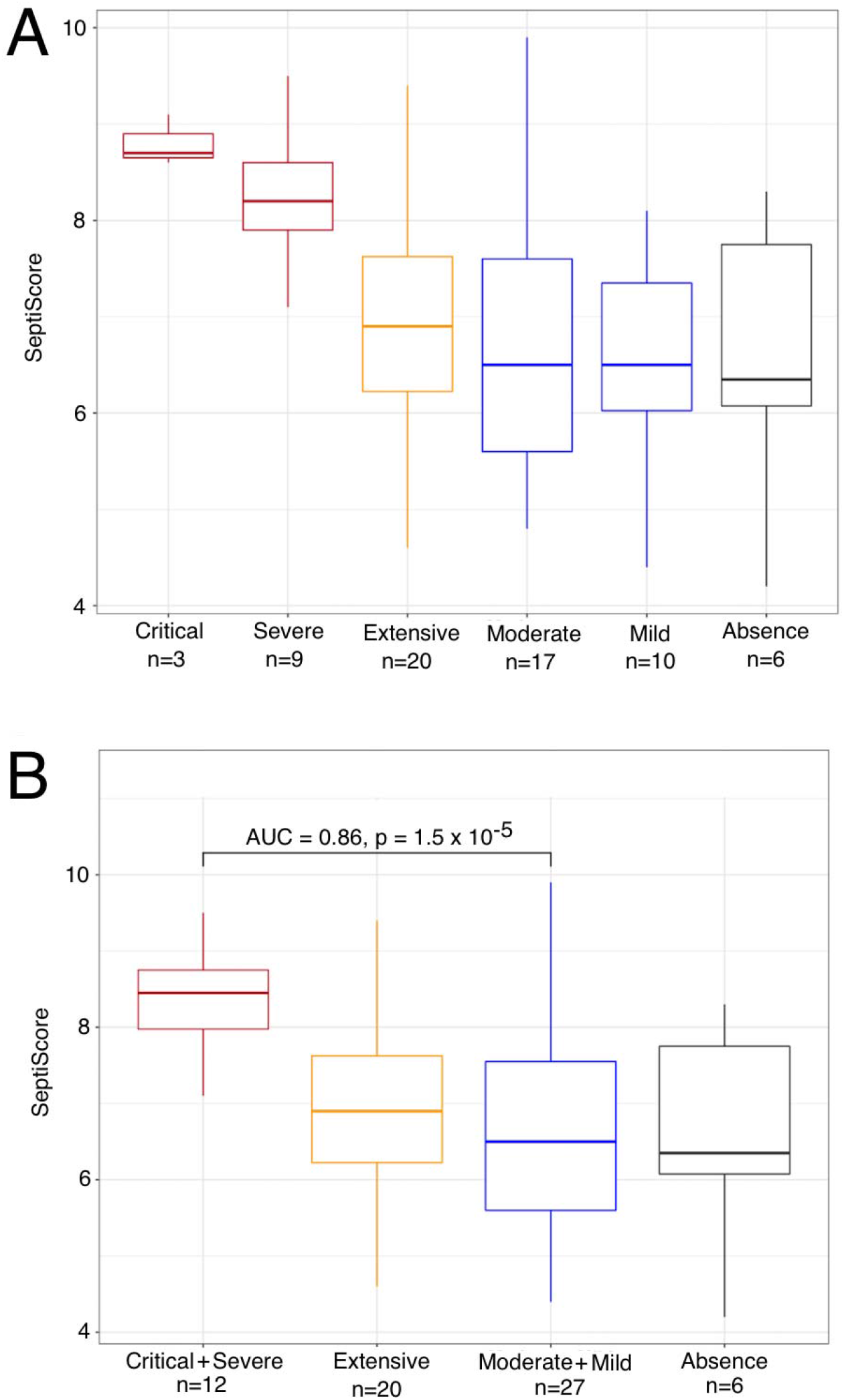
SeptiCyte RAPID scores, stratified by COVID-19 severity as measured by chest CT scans within 24 hours of SeptiCyte RAPID testing. (A) Stratification into critical, severe, extensive, moderate, mild, and absence groups based on chest CT scans. (B) Stratification with critical+severe combined, and moderate+mild combined.

## Results

The main characteristics of the patients in this study are summarized in Table 1. Obesity, characterized by a body mass index (BMI) > 30, was not reported in this table because this parameter was available only for 46 patients. Additional risk factors (including BMI when available) and treatments of the patients during their hospitalization are indicated in Supplementary Table 1. The distribution of SARS-CoV-2 (+) patients across COVID-19 severity categories and hospital locations is indicated in Supplementary Table 2.

**Table 1:** Main characteristics of COVID-19 patients in this study, according to the department where the sample for SeptiScore measurement was collected and patient follow-up occurred. Range is specified in [ ] square brackets, and percentage is specified in () parentheses. Abbreviation: NA, not applicable. ANOVA was used to calculate p-values.

|  | Emergency Department (fig. 1a) (n=67) |  |  |  |  | ICU<br>(fig. 1b) | Conven-<br>tional unit,<br>viremia<br>(fig. 1c) | Bacterial<br>Sepsis<br>(ICU) | p-value |
| --- | --- | --- | --- | --- | --- | --- | --- | --- | --- |
| Follow-up | Discharged<br>(< 24 hours<br>in ED) | Conventional<br>Hospital-<br>ization | Delayed<br>ICU<br>admission | Immediate<br>ICU<br>admission | Deceased<br>in ED |  |  |  |  |
| n | 25 | 33 | 4 | 3 | 2 | 23 | 4 | 8 |  |
| F/M | 15/10 | 15/18 | 2/2 | 1/2 | 2/0 | 8/15 | 1/3 | 2/6 | 0.4 |
| Median age (years) | 60<br>[20 – 84] | 72<br>[23 – 96] | 66<br>[63 – 66] | 76<br>[69 – 78] | 89<br>[85 – 93] | 57<br>[23 – 78] | 56<br>[38 – 67] | 62<br>[35 – 90] | 0.001 |
| Median time between<br>COVID-19 diagnosis and<br>SeptiCyte result, days<sup>1</sup> | 0<br>[0 - 12] | 0<br>[0 - 7] | 0.5<br>[0 - 16] | 0 | 0 | 3<br>[0 – 25] | 8.5<br>[1 – 10] | NA | <0.001 |
| Median time between<br>onset of symptoms and<br>SeptiCyte result, days<br>(symptomatic patients) | 5 (n=21)<br>[1 – 12] | 7 (n=31)<br>[0 – 30] | 6.5 (n=4)<br>[4 – 9] | 7 (n=3)<br>[4 – 10] | NA | 10.5<br>[4 – 42] | 14<br>[10 – 21] | 3<br>[1 - 10] | <0.001 |
| Median number of<br>comorbidities | 1 [0-4] | 2 [0-4] | 1.5 [1-3] | 2 [0-4] | 2 [1-3] | 3 [2-4] | 2 [1-2] | 2 [0-5] | 0.04 |
| CT-scan performed,<br>n (%) | 12 (48) | 30 (90.1) | 4 (100) | 2 (66.7) | 1 (50) | 15 (65.2) | 4 (100) | 0(0) | <0.001 |
| Alive, n (%) | 25 (100) | 27 (81.8) | 4 (100) | 3 (100) | 0 (0) | 15 (65.2) | 4 (100) | 7 (87.5) | <0.001 |
| Median length of<br>hospitalization, days | 0 (100) | 7<br>[2 – 22] | 21<br>[6 – 37] | 20<br>[7 – 38] | 1<br>[1-1] | 24<br>[6 – 74] | 10<br>[3 – 13] | 10<br>[6 – 34] | <0.001 |
| Median delay to death<br>(days)<sup>2</sup> | no cases | 8<br>[5 – 17] | no cases | no cases | 1<br>[1 - 1] | 24<br>[7 – 55] | no cases | 7<br>[7 - 7] | 0.005 |

Gravrand et al. (2022) Stratification of COVID-19 with SeptiCyte RAPID
|  | Emergency Department (fig. 1a) (n=67) |  |  |  |  | ICU<br>(fig. 1b) | Conven-<br>tional unit,<br>viremia<br>(fig. 1c) | Bacterial<br>Sepsis<br>(ICU) | p-value |
| --- | --- | --- | --- | --- | --- | --- | --- | --- | --- |
| Follow-up | Discharged<br>(< 24 hours<br>in ED) | Conventional<br>Hospital-<br>ization | Delayed<br>ICU<br>admission | Immediate<br>ICU<br>admission | Deceased<br>in ED |  |  |  |  |
| <sup>1</sup>Positive COVID diagnosis is equated with a (+) RT-qPCR test result for SARS-CoV-2.<br><sup>2</sup>Time between the first day of hospitalization and death. |  |  |  |  |  |  |  |  |  |

### Variations of SeptiScore and other biological markers according to patient group

Variations of the SeptiScore were analyzed in comparison to other well recognized biological markers of severity of SARS-CoV-2 infection (variations in leucocyte differential, including CD16-positive monocytes, CRP, D-dimer and lactate). As can be seen in Table 2 and Fig. 2 the SeptiScore increased with the severity of the disease. In considering the entire 94 patient cohort, using samples taken at admission, SeptiScore was significantly correlated with CRP (Pearson sample correlation coefficient ρ = 0.513; p < 0.0001), lactate (ρ = 0.321; p = 0.014) lymphocyte count (ρ = -0.354; p = 0.0008) and monocyte count (ρ = -0.219; p = 0.038).

**Table 2:** Comparison of the Septicyte score with conventional biological parameters and the hospital locations. Values expressed as median and [range]. Abbreviations: CRP, C-reactive protein; NA, not available. p-values calculated by one-way ANOVA, which used 102 out of 131 samples.

| Follow-up | Emergency Department (Fig 1a) |  |  |  |  | ICU (Fig 1b) | Conventional unit, viremia (Fig 1c) | Bacterial Sepsis (ICU) | p-value |
| --- | --- | --- | --- | --- | --- | --- | --- | --- | --- |
|  | Discharged (< 24 hours in ED) | Conventional Hospitalization | Delayed ICU admission | Immediate ICU admission | Deceased in ED |  |  |  |  |
| n | 25 | 33 | 4 | 3 | 2 | 23 | 4 | 8 |  |
| Septicyte score (A.U) | 6.4<br>[3.1 – 8.3] | 6.4<br>[3.5 – 9.9] | 8<br>[6.4 – 8.6] | 8.1<br>[7.7 – 8.2] | 7.2<br>[6.5 – 7.9] | 7.6<br>[5.3 – 9.5] | 6.2<br>[4.8 – 6.7] | 8.5<br>[2.9 – 11.1] | 0.003 |
| White blood cell (x 10<sup>9</sup>/L) | 5<br>[1.9 – 5.5] | 7.1<br>[2 – 24] | 5.3<br>[3.5 – 8.1] | 9.4<br>[7.5 – 10.7] | 12.6<br>[9.8 – 15.3] | 8.2<br>[2.2 – 39.7] | 6.3<br>[4.8 – 7.1] | 13.3<br>[4.4 – 24.3] | 0.020 |
| Platelets (x 10<sup>9</sup>/L) | 197<br>[97 – 708] | 198<br>[86 – 588] | 180<br>[117 – 311] | 316<br>[136 – 421] | 178<br>[139 – 217] | 238<br>[68 – 433] | 182<br>[167 – 242] | 180<br>[17 – 330] | 0.7 |
| Neutrophils (x 10<sup>9</sup>/L)) | 3.4<br>[1.1 – 11.6] | 5.5<br>[1.1 – 22.8] | 4.1<br>[2.2 – 5.7] | 6.8<br>[6.8 – 9.5] | 10.6<br>[8.6 – 12.7] | 6.8<br>[1.8 – 33.4] | 4.3<br>[3.6 – 6] | 8.6<br>[3.1 – 18.0] | 0.04 |
| Lymphocytes (x 10<sup>9</sup>/L) | 0.9<br>[0.4 – 3.6] | 0.8<br>[0.2 – 2.2] | 1.1<br>[0.4 – 1.4] | 0.7<br>[0.5 – 1.4] | 1.1<br>[0.5 – 1.6] | 0.65<br>[0.2 – 0.8] | 0.8<br>[0.7 – 1.7] | 1<br>[0.3 – 2.0] | 0.3 |
| Monocytes (x 10<sup>9</sup>/L) | 0.6<br>[0 – 1.2] | 0.6<br>[0.1 – 1.5] | 0.4<br>[0.2 – 1.2] | 0.5<br>[0.2 – 1.1] | 0.8<br>[0.6 – 0.9] | 0.5<br>[0.1 – 2.1] | 0.6<br>[0.3 – 0.9] | 0.8<br>[0.1 – 1.4] | >0.9 |
| CD16<sup>POS</sup> monocytes (x 10<sup>9</sup>/L) | 0.07<br>[0 – 0.38] | 0.059<br>[0.003 – 0.29] | 0.033<br>[0.022 – 0.058] | 0.053<br>[0.005 – 0.094] | 0.064 | 0.020<br>[0.005 – 0.12] | 0.058<br>[0.044 – 0.074] | 0.094<br>[0.004 – 0.16] | 0.7 |
| CRP (mg/L) | 22<br>[1 – 279] | 95<br>[1 – 287] | 166<br>[84 – 181] | 173<br>[105 – 296] | 111<br>[86 – 137] | 161<br>[23 – | 47<br>[2 – 154] | 196<br>(159-233) | <0.001 |

Gravrand et al. (2022) Stratification of COVID-19 with SeptiCyt RAPID
|  | Emergency Department (Fig 1a) |  |  |  |  | ICU<br>(Fig 1b) | Conventional<br>unit, viremia<br>(Fig 1c) | Bacterial<br>Sepsis (ICU) | p-value |
| --- | --- | --- | --- | --- | --- | --- | --- | --- | --- |
| Follow-up | Discharged<br>($< 24$ hours<br>in ED) | Conventional<br>Hospital-<br>ization | Delayed ICU<br>admission | Immediate<br>ICU<br>admission | Deceased<br>in ED | | | | |
|  |  |  |  |  |  | 270] |  |  |  |
| D-dimer (mg/L) | 0.79<br>[0.22 – 3.80] | 0.66<br>[0.28 – 2.21] | 1.07<br>[0.43 –<br>92.86] | 1.59<br>[0.54 – 1.81] | 6.961 | 0.95<br>[0.43 –<br>16.25] | 1.29<br>[0.83 – 1.74] | NA | 0.02 |
| Lactate<br>(mmol/L) | 0.8<br>[0.6 – 1.2] | 1.1<br>[0.9 – 5.3] | 1.6<br>[1.5 – 1.7] | 2.6<br>[1.1 – 2.6] | 4.2<br>[3.2 – 5.1] | 1.5<br>[0.8 – 2.5] | 1.8 | 1.3<br>(0.6-8.0) | 0.01 |

SeptiScores were significantly higher in patients requiring ICU hospitalization, compared to patients who were immediately discharged or transferred to conventional units (p = 0.0004, Kruskal-Wallis test). SeptiScores for the ICU patients were similar in value to scores for 8 SARS-CoV-2 (-) patients having culture-confirmed bacterial sepsis (Fig. 2).

A receiver operating characteristic (ROC) curve analysis was performed on the different biological parameters recorded during the study, in order to identify patients requiring ICU hospitalization. An AUC > 0.7 was considered to be relevant as a discriminating parameter [16]. Using data from the earliest measured values of the parameters, only two biological parameters fulfilled this criteria, SeptiScore and CRP, with an AUC of 0.78 and 0.79, and a binary cut-off of 6.6 and 100 mg/L, respectively.

### Variation in SeptiScore, according to lung injury observed in CT-scan

For 68 patients, a CT-scan was performed within 24 hours after admission to the hospital. Three of the patients had non-specific presentations in CT-scan, and were not considered further. Six had an absence of lung damage, while the remaining patients could be classified as having mild (N=10), moderate (N=17), extensive (N=20), severe (N=9) or critical (N=3) extents of lung damage. As can be seen in Fig. 3, the SeptiScore increased with the extent of the lung injury, the highest SeptiScore values being observed in patients with severe or critical lung injuries. Following from this analysis, patients were divided in two groups: those with severe or critical (≥ 50%) lung injury (n = 12), vs. those with specific but <50% lung injury (n = 53) as defined by chest CT scan. ROC curve analysis gave AUC = 0.86, with a SeptiScore cut-off of 7 providing a good discrimination between these two groups. CRP, lymphocytes, monocytes, CD16-positive monocytes and lactate were also discriminatory for patients with ≥ 50% versus <50% lung injury (Table 3).

**Table 3:** AUC and cut-off values of the different biological parameters tested for the discrimination of patients with lung injury <50% (absence+mild+moderate+extensive) versus ≥50 % (severe+critical) on initial CT-scan. The first measured timepoint data were used for parameter values. Abbreviations: AUC, area under the curve; CRP, C-reactive protein.

|  | N, lower damage (<50%) | N, higher damage (≥50%) | p-value (t-test) | AUC | Cut-off | Sensitivity (%) | Specificity (%) |
| --- | --- | --- | --- | --- | --- | --- | --- |
| SeptiScore | 53 | 12 | 0.00013 | 0.860 | ≥ 7 | 100.0 | 61.1 |
| CRP (mg/L) | 49 | 11 | 0.00032 | 0.836 | > 154.1 | 81.8 | 78.4 |
| Lymphocytes (10<sup>9</sup>/L) | 52 | 11 | 0.025 | 0.786 | ≤ 0.7 | 81.8 | 62.3 |
| Monocytes (10<sup>9</sup>/L) | 52 | 11 | 0.016 | 0.758 | ≤ 0.5 | 81.8 | 56.6 |
| Monocytes CD16<sup>POS</sup> (10<sup>9</sup>/L) | 43 | 10 | 0.046 | 0.773 | ≤ 0.037 | 80 | 66.7 |
| Lactate (mmol/L) | 35 | 11 | 0.114 | 0.745 | ≥ 1.2 | 81.8 | 66.7 |

Interestingly, SeptiScores and CT scans were determined three times for two patients while they were in the ICU. Each time, both patients had lung injury ≥ 50% and their median SeptiScore was 8.4 (range 6 to 9.7), while their median CRP had decreased to 2.5 mg/L presumably because of their treatments with corticosteroids and tocilizumab.

### Elevated SeptiScore is predictive of ICU admission in patients with extensive lung injury

All the patients except for one with lung injury ≥ 50 % were admitted to ICU. In addition, among the 17 patients with extensive lung injury (between 25 – 50 %) whose age was less than 80 years, 6 were admitted to ICU, either immediately after hospital admission (3 cases) or after some days in conventional units (3 cases). For patients with extensive lung injury and age <80 years, SeptiScores were significantly higher in those requiring ICU care (N=6) versus those who did not require this level of care (N=11), with median SeptiScore values of 7.6 versus 6.6, respectively (p = 0.014 by Welch’s two sample t-test). In contrast, CRP levels were not significantly different for these patients (median levels 83 versus 114 mg/L, respectively (p = 0.71 by Welch’s two sample t-test). The SeptiScore AUC was 0.86, and an optimal SeptiScore cut-off for this discrimination was 6.7.

### SeptiCyte RAPID, compared to / combined with other clinical variables

We conducted ROC curve analyses to compare SeptiCyte RAPID to other single biomarkers, specifically CRP, D-dimer, lactate, monocytes, and CD16-monocytes, for discriminating (critical + severe) vs. (moderate + mild) cases. These biomarkers have all been proposed to be used by clinicians and investigators to assess the severity of COVID-19 clinical trajectories.

In these analyses, we compared critical + severe cases in ICU (n=11) versus moderate + mild cases not in ICU (n=24), with only the first SeptiCyte RAPID measurement considered. The complete combinatorial analysis is presented in Supplementary Table 3. We found that the mean AUC for SeptiCyte RAPID was greater than that of any individual biomarker and of most combinations thereof (Fig. 4A).

**Figure 4.**
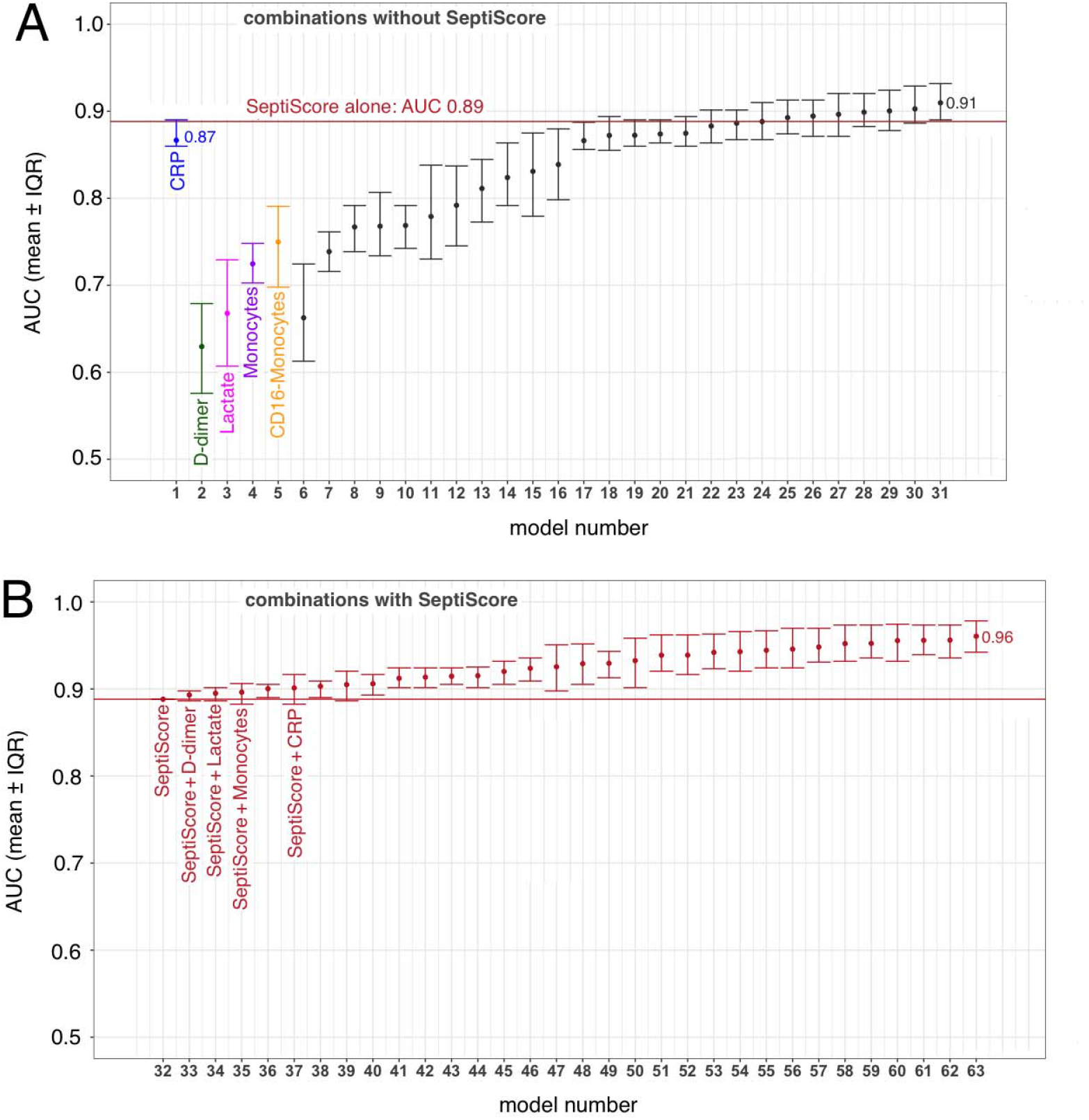
Combining SeptiScore with other clinical variables. The variables CRP, D-dimer, lactate, monocytes, CD16-monocytes were examined alone and in combination. The patients included in this analysis were as follows: Critical + Severe in ICU (n=11) versus Moderate + Mild not in ICU (n=24), with only the first SeptiCyte RAPID measurement considered. The identities of the regression models are given in Supplementary Table 3. Points indicate mean AUC values, and error bars indicate the interquartile range of AUCs for 100 imputation replicates. (A) Combinations without SeptiScore. (B) Combinations with SeptiScore.

The single biomarker that came closest to SeptiCyte RAPID in performance was CRP (AUC 0.87, versus SeptiScore AUC 0.89). In this regard, however, we note that SeptiCyte RAPID remains discriminatory (i.e. sensitive to lung injury) during the follow-up period, when CRP has been artificially reduced due to treatment with corticosteroids and tocilizumab (see Discussion).

Combining the SeptiScore with one or more of these other biomarkers provided an additional improvement in AUC beyond the use of the SeptiScore alone (Fig. 4B). Any of the logistic regression models containing SeptiScore with clinical parameters (Panel B) performed better than all of the singleton biomarker models, and better than most models that did not contain SeptiScore (compare panels B and A).

### Longitudinal monitoring of patient trajectories

Finally, we consider the use of SeptiCyte RAPID, as a component of the longitudinal monitoring of COVID-19 patients during their transit through hospital. Fig. 5 presents the clinical trajectory of one such patient, who presented to ED, was transferred immediately to ICU, suffered a critical level of lung damage (>75%), received intubation with mechanical ventilation, and unfortunately died. An important point to note about this patient’s trajectory is that an initially high CRP level (112.5 mg/L) was greatly reduced (to 2.5 mg/L) by tocilizumab + corticosteroids. However the SeptiScore, initially 9.1, remained high throughout the patient’s trajectory (8.1, 7.0, 7.1, 8.8 upon repeat testing).

**Figure 5.**
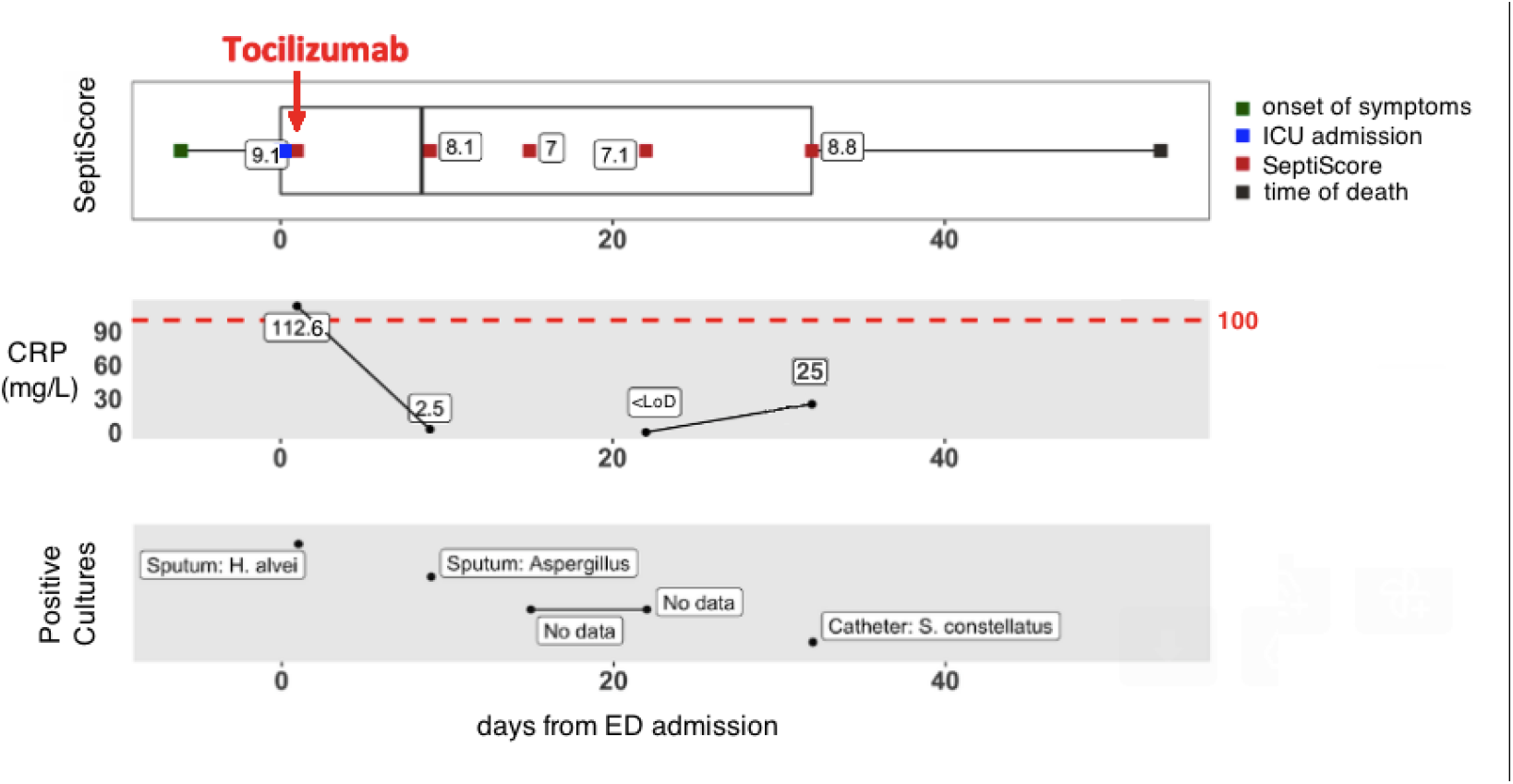
Hospital trajectory of patient with “critical” lung damage (male, between 60-70 years old, >75% lung injury (critical), intubated with mechanical ventilation, died). In the CRP panel, the limit of detection (LoD) = 1 mg/L).

## Discussion

Infection by SARS-CoV-2, initially described in China in 2019, rapidly spread throughout the world and has evolved in several waves, corresponding to the appearance of new variants. These waves are characterized by the influx of patients to emergency departments, which are quickly saturated. Therefore, it is important to have a rapid diagnosis which identifies patients with mild forms of the disease who can immediately be discharged, versus those who require hospitalization, or even admission to intensive care.

In addition, identification of potential risk factors that could potentially predict the disease course may be of great utility for healthcare professionals to efficiently triage patients, personalize treatment, monitor clinical progress, and allocate proper resources at all levels of care to mitigate morbidity and mortality. Different risk factors, which can be divided in demographic, clinical, hematological, biochemical, and radiographic factors, have been so far described to evaluate COVID-19 disease severity [6]. Although certain factors clearly identify patients at high risk, none can be considered a gold standard.

At this time, in addition to the clinical examination and hypoxemia, evaluation of disease severity is based on hematological tests (severity of the lymphopenia is considered as a risk factor), biochemical tests to evaluate the inflammatory state (mainly CRP), and CT-scans [17], with the patient’s age and comorbidities also taken into account. However, hospital central laboratories and imaging departments may be quickly overwhelmed by the importance of the requests, potentially leadinng to delays in the return of results, and consequently to emergency department overcrowding. Therefore it is important to have tests allowing a rapid assessment of the severity of the infection of patients.

In this study we have evaluated the performance of a new host immune response test, SeptiCyte RAPID, in discriminating COVID-19 severity in confirmed SARS-CoV-2 positive patients who were admitted to the ED. The study cohort reflected the real-world situation at this stage of the pandemic: it consisted of a heterogeneous group of non-acute and acutely ill patients with varying phases of an inflammatory response as well as a range of comorbidities.

SeptiCyte RAPID was found to have utility for diagnosing severe or critical COVID-19 and the subsequent risk of ICU admission in SARS-CoV-2 positive patients. At time of presentation, SeptiCyte RAPID differentiated critical+severe vs. moderate+mild COVID-19 with AUC 0.86 (Figure 3B). If the critical+severe cohort was restricted to those measured in ICU vs. moderate+mild measured outside ICU then the AUC increases to 0.89 (Figure 4). In patients requiring ICU hospitalization, the SeptiScore was similar to values observed in patients with culture-confirmed bacterial sepsis.

Additionally, the SeptiScore discriminated patients with lung injury above 50% on CT-scan (AUC = 0.88 for critical+severe vs. absence+mild+moderate+extensive), and particularly could identify patients with extensive lung injury (25 – 50%) requiring ICU admission. Lastly, during the follow-up of the patients, the SeptiScore remained elevated in patients will lung injury above 50%, whereas other biological markers such as CRP decreased, likely due to anti-inflammatory treatments received by the patients (corticosteroids, tocilizumab).

We note that SeptiCyte RAPID was initially validated for use in patients exhibiting ≥2 SIRS criteria and suspected of sepsis, with bacterial infection being the most common etiology [8]. Optimal application of the test for discriminating COVID-19 severities, or for guiding the ICU admission decision for COVID-19 patients, could involve minor adjustment to the predefined assay cutoffs used by the SeptiCyte RAPID software. Testing of additional COVID-19 patients may provide further insight on this point.

In comparing SeptiCyte and CRP, several points should be further emphasized. The study cohort was heavily treated with both corticosteroids (59/94 = 63%) and tocilizumab (24/94 = 26%), with some patients receiving both therapies (22/94 = 23%). It is well known that CRP is negatively modulated by both corticosteroids and tocilizumab [18-23]. Thus, in treated patients, CRP may no longer be a useful indicator of progression to more severe states, e.g. secondary bacterial infections [20, 24] whereas SeptiScore values were not impacted, consistent with its original function as an indicator of sepsis. The patient trajectory described in Fig. 5 illustrates this point: an initially high CRP level was greatly reduced by tocilizumab + corticosteroids, but the SeptiScore remained high throughout the patient’s trajectory. In this particular case, while CRP may have been an accurate indicator of inflammatory state, it clearly was unable to indicate the underlying condition that eventually led to the patient’s death. In contrast, the SeptiScore remained high and at an alert level.

Another point to note is that CRP is an integral component of the “acute phase response” which is an early and relatively non-specific part of the innate immune response to various stressors including trauma and infection by both bacteria and viruses [25, 26]. The two genes of SeptiCyte RAPID (PLAC8 and PLA2G7) are not in the acute phase response pathway, so it is perhaps unsurprising that they behave differently in response to anti-inflammatory treatments.

One strength of our study is that test results were returned quickly in real time, typically within 1-2 hours of sample collection, in a near-patient setting (even in the ED). An additional strength of the study is that it is representative of the real-world situation that occurred in our institution over the study period. However, it is clear that is also a limitation, since the patient population was quite heterogeneous in terms of pre-existing conditions and comorbidities. This study design limitation could lead to multiple variables contributing to an elevation of the SeptiCyte RAPID score. A further limitation is that most of the patients admitted to ICU had blood drawn after ICU admission. Consequently, the SeptiScore was perhaps different (possibly higher) when patients were in the ED. This question will need to be addressed in subsequent studies.

## Conclusions

This study supports the potential use of SeptiCyte RAPID in the risk stratification of COVID-19 patients, as measured by clinical severity assessment based on chest CT scans and/or ICU admission. With a quick availability from whole blood (potentially within 1 hour) and with minimal user interaction, the assay may be helpful in stratifying or triaging patients during their transit through hospital. In contrast, hematological and biochemical markers as well as CT scans can take several hours, especially in the event of a massive influx of patients to the hospital. Secondly, small hospitals do not always have an imaging department available 24 hours a day, and the SeptiScore result gives good information in the event of lung damage greater than 50%. Thirdly, it can be useful for the follow-up of patients, particularly patients in the ICU, who are difficult to transport to the imaging department, and for whom the interventional treatments may have normalized the usual biochemical and/or hematological parameters (such as CRP). The SeptiCyte RAPID test could also constitute an approach to evaluate lung damages. Moreover, the evolution of patients with COVID-19 in intensive care is often marked by the appearance of bacterial complications, for which the SeptiScore has shown potential value (Fig. 5).

## Supporting information

Strobe Checklist

## Data Availability

All data produced in the present study are available upon reasonable request to the authors

## Abbreviations

AHT: arterial hypertension
ARDS: acute respiratory distress syndrome
AUC: area under ROC curve
CD16: Fc receptor with low affinity for IgG, encoded by genes FCGR3A and FCGR3B
BMI: body mass index
CI: confidence interval
COVID-19: coronavirus disease 2019
CRP: C-reactive protein
CT: computed tomography
ECMO: extracorporeal membrane oxygenation
ED: Emergency Department
EDTA: ethylene diamine tetraacetic acid
F: female
GGO: ground glass opacity
ICU: Intensive Care Unit
M: male
NIH: United States National Institutes of Health
NGS: next-generation sequencing
NA: not available
PCT: procalcitonin
PLA2G7: Phospholipase A2 Group VII gene
PLAC8: Placenta Associated 8 gene
ROC: receiver operating characteristic [curve]
RT-qPCR: reverse transcription - quantitative polymerase chain reaction
SARS-CoV-2: severe acute respiratory syndrome coronavirus 2
SPC: sample processing control
WHO: World Health Organization

## Acknowledgements

We thank Biocartis for provision of the Idylla platform to the Foch Hospital site, and Julie Gaiffas and David Favy of Biocartis for liason support. This project has been supported in part by contract # 75A50120C00125 from the DRIVe Solving Sepsis program of the Biomedical Advanced Research and Development Authority (BARDA), part of the US HHS Office of the Assistant Secretary for Preparedness and Response. We also thank Immunexpress for continued financial support over the course of this project. We thank Dr. Dayle Sampson for initial R coding for analysis leading to Fig. 4.

## Contributions

Conceptualization: MV

Methodology: MV, VG, JTK

Investigation: MV

Supervision: MV

Data acquisition, VG, FM, FA, MCB, BZ, FP, TF, EF, MV

Data curation: MV, KN

Custom software: KN

Visualization: KN, TDY, MV

Formal analysis: MV, VG, KN, TDY

Writing—initial draft preparation: MV, TDY

Writing—review and editing: MV, BZ, FM, TP, EF,TDY, KN, JTK

Funding acquisition: MV, TDY

All authors have read and agreed to the published version of the manuscript.

## Data availability statement

Data will be provided, upon reasonable request to corresponding author.

## Corresponding author

Correspondence to Marc Vasse at email address

