## Supplementary Tables for "Stratification of COVID-19 severity using SeptiCyte RAPID, a novel host immune response test"

Supplementary Table 1: Patient comorbidities and treatments

| Characteristic | Stratum | N (%) |
| --- | --- | --- |
| Number of comorbidities | 5 | 1 (1.0%) |
|  | 4 | 5 (5.3%) |
|  | 3 | 13 (13.8%) |
|  | 2 | 30 (31.9 %) |
|  | 1 | 23 (24.5.0%) |
|  | 0 | 22 (23.4 %) |
| Types of  comorbidities | None listed | 22 (23.4%) |
|  | Hypertension | 42 (44.7%) |
|  | Diabetes | 32 (34.0 %) |
|  | Obesity (BMI ≥ 30)* | 22 (23.4%) |
|  | Pulmonary | 15 (16.0%) |
|  | Renal | 10 (10.6%) |
|  | Cardiovascular | 13 (13.8 %) |
|  | Cancer | 10 (10.6%) |
|  | Dyslipidemia | 6 (6.4 %) |
|  | Hypothyroidism | 5 (5.3%) |
| Treatments | ICU | 30 (31.9%) |
|  | Corticosteroids | 51 (54.3%) |
|  | Tocilizumab | 24 (25.5 %) |
|  | Oxygen | 57 (60.6%) |
|  | Intubation | 7 (7.4%) |
|  | ECMO | 3 (3.2%) |
|  | Antibiotics | 36 (38.3%) |

*BMI available for only 46 patients

Supplementary Table 2 : Distribution of SARS-CoV-2 (+) patients across COVID-19 severity categories and hospital locations. COVID-19 severity assessments were made on the basis of chest CT scans, conducted within 24 hours of SeptiCyte RAPID measurement. Range is specified in [ ] square brackets, and percentage is specified in ( ) parentheses. The median values for age and the median time between onset of symptoms and SeptiCyte result are reported for those patients with CT scan results.

|  | Emergency Department (Fig 1a) (n=67 total; 49 with CT scan results) | | | | | ICU  (Fig 1b) | Conventional Unit,Viremia  (Fig. 1c) |
| --- | --- | --- | --- | --- | --- | --- | --- |
| Follow-up | Discharged  (< 24 hours  in ED) | Conventional  Hospitalization | Delayed ICU admission | Immediate ICU admission | Deceased in ED |  |  |
| n (total) = 94 | 25 | 33 | 4 | 3 | 2 | 23 | 4 |
| n (with CT scan result) | 12 | 30 | 4 | 2 | 1 | 15 | 4 |
| F/M (with CT scan result) | 6/6 | 13/17 | 2/2 | 0/2 | 1/0 | 6/9 | 1/3 |
| Median age, years | 58  [20 – 84] | 71  [23 – 96] | 66  [63 – 66] | 77  [76 – 78] | >80 | 59  [27 – 78] | 57  [38 – 67] |
| Median time between onset of symptoms and  SeptiCyte result, days (symptomatic patients) | 7 (n=11)  [1 – 12] | 7 (n=28)  [0 – 30] | 6 (n=4)  [4 – 9] | 8.5 (n=2)  [7 – 10] | NA | 10  [1 – 24] | 9  [9 – 11] |
| Radiologic injury, n (%) |  |  |  |  |  |  |  |
| Absence (0%) | 5 (20%) | 1 (3%) | 0 | 0 | 0 | 0 | 0 |
| Mild (present but <10 %) | 1 (4%) | 8 (24%) | 0 | 0 | 0 | 0 | 1 (25%) |
| Moderate [10 – 25 %] | 4 (16%) | 8 (24%) | 1 (25%) | 0 | 1 (50%) | 2 (8.7%) | 1 (25%) |
| Extensive (25 - 50 %) | 1 (4%) | 11 (33%) | 1 (25%) | 1 (33.3%) | 0 | 4 (17%) | 2 (50%) |
| Severe [50 – 75 %] | 0 | 1 (3%) | 1 (25%) | 1 (33.3%) | 0 | 6 (26%) | 0 |
| Critical (> 75 %) | 0 | 0 | 0 | 0 | 0 | 3 (13.1%) | 0 |
| Not specific | 1 (4%) | 1 (3%) | 1 (25%) | 0 | 0 | 0 | 0 |
| Not performed | 13 (52%) | 3 (9.1%) | 0 | 1 (33.3%) | 1 (50%) | 8 (34.8%) | 0 |

Supplementary Table 3: Logistic Regression models (see Figs. 4A, 4B in main text). A complete permutation analysis of different logistic combinations of up to six variables (SeptiScore, monocytes, CD16-monocytes, D-dimer, lactate and CRP) was conducted, resulting in 63 independent logistic regression models. The models were used to discriminate Critical + Severe in ICU (n=13) vs. Moderate + Mild not in ICU (n = 24) and their AUC values were calculated. Interquartile ranges for AUC were obtained by resampling from 100 replicate datasets. Missing values in the line data were imputed using Amelia, a multiple imputation algorithm [1]. Multiple imputation is claimed to have reduced bias and increased accuracy over point-imputation methods such as "mean" or "median" imputation [1]. (The numbers of missing values were 1 for monocytes; 7 for CD16-monocytes, 9 for D-dimer; 11 for lactate; and 1 for CRP.) Abbreviations: CRP, C-reactive protein; Mono, monocytes; Mono.CD16, CD16-monocytes.

| **Model** | **variables** | **MEAN** | **MEDIAN** | **25%** | **75%** |
| --- | --- | --- | --- | --- | --- |
| 1 | CRP | 0.867 | 0.871 | 0.860 | 0.890 |
| 2 | D-dimer | 0.630 | 0.640 | 0.576 | 0.679 |
| 3 | Lactate | 0.668 | 0.680 | 0.607 | 0.729 |
| 4 | Mono | 0.725 | 0.722 | 0.703 | 0.748 |
| 5 | Mono.CD16 | 0.750 | 0.752 | 0.698 | 0.791 |
| 6 | D-dimer + Lactate | 0.663 | 0.674 | 0.613 | 0.724 |
| 7 | Lactate + Mono | 0.739 | 0.742 | 0.716 | 0.761 |
| 8 | D-dimer + Lactate + Mono | 0.767 | 0.765 | 0.739 | 0.792 |
| 9 | Mono + Mono.CD16 | 0.768 | 0.758 | 0.734 | 0.807 |
| 10 | D-dimer + Mono | 0.769 | 0.769 | 0.742 | 0.792 |
| 11 | Lactate + Mono.CD16 | 0.779 | 0.778 | 0.730 | 0.838 |
| 12 | Lactate + Mono + Mono.CD16 | 0.792 | 0.780 | 0.745 | 0.837 |
| 13 | D-dimer + Mono.CD16 | 0.811 | 0.811 | 0.773 | 0.845 |
| 14 | D-dimer + Mono + Mono.CD16 | 0.824 | 0.822 | 0.792 | 0.864 |
| 15 | D-dimer + Lactate + Mono.CD16 | 0.831 | 0.831 | 0.779 | 0.875 |
| 16 | D-dimer + Lactate + Mono + Mono.CD16 | 0.839 | 0.841 | 0.798 | 0.880 |
| 17 | CRP + D-dimer | 0.866 | 0.867 | 0.856 | 0.887 |
| 18 | CRP + Mono | 0.872 | 0.883 | 0.855 | 0.894 |
| 19 | CRP + Lactate | 0.872 | 0.875 | 0.860 | 0.890 |
| 20 | CRP + D-dimer + Lactate | 0.874 | 0.875 | 0.864 | 0.890 |
| 21 | CRP + Lactate + Mono | 0.875 | 0.883 | 0.860 | 0.894 |
| 22 | CRP + D-dimer + Mono | 0.883 | 0.886 | 0.864 | 0.902 |
| 23 | CRP + D-dimer + Lactate + Mono | 0.886 | 0.888 | 0.867 | 0.902 |
| 24 | CRP + Mono.CD16 | 0.888 | 0.890 | 0.867 | 0.910 |
| 25 | CRP + Mono + Mono.CD16 | 0.893 | 0.890 | 0.874 | 0.913 |
| 26 | CRP + Lactate + Mono.CD16 | 0.895 | 0.894 | 0.871 | 0.913 |
| 27 | CRP + D-dimer + Mono.CD16 | 0.896 | 0.896 | 0.871 | 0.920 |
| 28 | CRP + Lactate + Mono + Mono.CD16 | 0.899 | 0.898 | 0.883 | 0.920 |
| 29 | CRP + D-dimer + Lactate + Mono.CD16 | 0.900 | 0.898 | 0.878 | 0.924 |
| 30 | CRP + D-dimer + Mono + Mono.CD16 | 0.903 | 0.905 | 0.886 | 0.929 |
| 31 | CRP + D-dimer + Lactate + Mono + Mono.CD16 | 0.910 | 0.909 | 0.890 | 0.932 |
| 32 | SeptiScore | 0.888 | 0.888 | 0.888 | 0.888 |
| 33 | SeptiScore + D-dimer | 0.893 | 0.890 | 0.886 | 0.898 |
| 34 | SeptiScore + Lactate | 0.895 | 0.894 | 0.886 | 0.902 |
| 35 | SeptiScore + Mono | 0.896 | 0.898 | 0.883 | 0.906 |
| 36 | SeptiScore + D-dimer + Mono | 0.900 | 0.900 | 0.890 | 0.905 |
| 37 | SeptiScore + CRP | 0.901 | 0.913 | 0.883 | 0.917 |
| 38 | SeptiScore + D-dimer + Lactate | 0.903 | 0.898 | 0.890 | 0.909 |
| 39 | SeptiScore + CRP + Lactate | 0.905 | 0.911 | 0.886 | 0.920 |
| 40 | SeptiScore + Lactate + Mono | 0.906 | 0.909 | 0.893 | 0.917 |
| 41 | SeptiScore + CRP + D-dimer | 0.912 | 0.917 | 0.902 | 0.924 |
| 42 | SeptiScore + D-dimer + Lactate + Mono | 0.914 | 0.909 | 0.902 | 0.924 |
| 43 | SeptiScore + CRP + Mono | 0.915 | 0.913 | 0.905 | 0.924 |
| 44 | SeptiScore + CRP + D-dimer + Lactate | 0.915 | 0.920 | 0.902 | 0.925 |
| 45 | SeptiScore + CRP + Lactate + Mono | 0.920 | 0.924 | 0.905 | 0.932 |
| 46 | SeptiScore + CRP + D-dimer + Mono | 0.924 | 0.922 | 0.909 | 0.936 |
| 47 | SeptiScore + Mono.CD16 | 0.925 | 0.920 | 0.898 | 0.951 |
| 48 | SeptiScore + Mono + Mono.CD16 | 0.929 | 0.924 | 0.905 | 0.952 |
| 49 | SeptiScore + CRP + D-dimer + Lactate + Mono | 0.930 | 0.932 | 0.913 | 0.943 |
| 50 | SeptiScore + Lactate + Mono.CD16 | 0.933 | 0.932 | 0.902 | 0.958 |
| 51 | SeptiScore + CRP + Mono.CD16 | 0.939 | 0.939 | 0.920 | 0.962 |
| 52 | SeptiScore + Lactate + Mono + Mono.CD16 | 0.939 | 0.943 | 0.917 | 0.962 |
| 53 | SeptiScore + D-dimer + Mono.CD16 | 0.942 | 0.943 | 0.923 | 0.963 |
| 54 | SeptiScore + CRP + Lactate + Mono.CD16 | 0.943 | 0.943 | 0.920 | 0.966 |
| 55 | SeptiScore + CRP + Mono + Mono.CD16 | 0.944 | 0.943 | 0.924 | 0.967 |
| 56 | SeptiScore + D-dimer + Mono + Mono.CD16 | 0.946 | 0.947 | 0.924 | 0.970 |
| 57 | SeptiScore + CRP + Lactate + Mono + Mono.CD16 | 0.948 | 0.947 | 0.931 | 0.970 |
| 58 | SeptiScore + D-dimer + Lactate + Mono.CD16 | 0.952 | 0.955 | 0.932 | 0.973 |
| 59 | SeptiScore + CRP + D-dimer + Mono.CD16 | 0.952 | 0.951 | 0.936 | 0.973 |
| 60 | SeptiScore + D-dimer + Lactate + Mono + Mono.CD16 | 0.956 | 0.955 | 0.932 | 0.974 |
| 61 | SeptiScore + CRP + D-dimer + Lactate + Mono.CD16 | 0.956 | 0.955 | 0.939 | 0.973 |
| 62 | SeptiScore + CRP + D-dimer + Mono + Mono.CD16 | 0.956 | 0.955 | 0.936 | 0.973 |
| 63 | SeptiScore + CRP + D-dimer + Lactate + Mono + Mono.CD16 | 0.961 | 0.962 | 0.942 | 0.978 |
